# Real-World Data Collection from Expanded Access Case Studies for the Treatment of Nontuberculous Mycobacterial Infection with Clofazimine

**DOI:** 10.1101/2023.10.30.23297757

**Authors:** Misty Gravelin, Theophilus Nguyen, Madeleine Davies, Blair Richards, Jonathan Z. Sexton, Kevin Gregg, Kevin J. Weatherwax

## Abstract

**Background:** Due to its indolent nature, nontuberculous mycobacteria (NTM) are increasing in global prevalence as a cause of pulmonary infections and are difficult to treat with traditional antibiotics. Here, we study the repurposing of clofazimine (CFZ) to treat NTM through expanded access in a single health system. Our main objectives are to describe the feasibility of accessing and analyzing expanded access data and to generate hypotheses regarding CFZ use in NTM treatment.

**Methods:** A retrospective chart review was performed on patients within a single health system who had been approved for expanded access of clofazimine or who received it through an outside hospital for NTM treatment. Data were collected on patients’ baseline demographics, details of their NTM infection, concomitant therapies, and results as of 30 June 2021.

**Results:** A total of 55 patients were identified upon initial review as potentially receiving CFZ for NTM infection. After excluding 19 patients who did not initiate CFZ, data from the remaining 36 patients were collected and summarized. The median age at which patients were diagnosed with NTM was 51.3 years old, with a median BMI of 21.2 kg/m^2^. Patients were more likely to be female (64%), have a baseline lung disease (72%), and 52% were current or former smokers at the time of their diagnosis. The most common species isolated was *M. avium* complex (47%) followed by *M. abscessus* (36%), with the most common site of infection being the lung (78%). The majority of patients presented with productive cough with excess sputum production followed by pulmonary nodules and bronchiectasis present on radiograph.

**Conclusions:** This study demonstrated the difficulty of collecting retrospective real-world data via electronic healthcare records on symptoms, side effects, and radiography from patients who obtained a drug through expanded access. Based on the findings of this study, we recommend further research into the potential use of CFZ in patients with *M. abscessus* pulmonary infections.

## Introduction

Over the past few decades, it has been confirmed that the main sources of nontuberculous mycobacteria (NTM) infecting humans and other species are from natural and human-engineered environments.^1-2^ NTM, which are opportunistic pathogens mainly for humans, are comprised of an ever-growing collection of mycobacterial species and subspecies that are ubiquitous in the environment.^3^ Similar to mycobacterium tuberculosis (MTB), NTM can cause persistent pulmonary diseases in patients, with increased risk in those with compromised immune systems or pulmonary disease. While NTM can cause extrapulmonary disease, such manifestations are less frequent.^4^

In the United States, the Centers for Disease Control and Prevention do not mandate reporting of NTM infections; hence, a national estimate of NTM epidemiology relies solely upon published scientific reports. One report of NTM pulmonary disease based on diagnosis codes to a single managed care company estimated the incidence of NTM in 2015 to be 11.7 per 100,000 person-years, a 7.5% increase from 2007.^5^ In comparison, a study of veterans with chronic obstructive pulmonary disease (COPD) estimated an increased NTM isolation incidence of 151.1% from 2011 to 2015.^6^ The incidence of NTM has been reported to increase not only in the United States but also in many Asian and European countries, and they have become a more common finding than the classical MTB in these countries.^7^ The slow-growing *M. avium* complex (MAC) have been identified to be the most common perpetrators of NTM-related disease in North America and Europe.^8^

Immediately after inhalation, NTM surface glycopeptidolipids bind to alveolar macrophage complement receptors.^9^ This interaction results in the internalization of the invading NTM. However, several NTM species have developed mechanisms to evade destruction by macrophages. The first of these strategies is the production of biofilms. One study indicated that when *M. avium* subspecies *hominissuis* is surrounded by a biofilm, it both evades destruction by macrophage and induces macrophage apoptosis.^10^ In addition to biofilm production, species of NTM such as *M. abscessus* have demonstrated the ability to halt phagosome maturation and acidification.^11^ Given the relatively slow growth of NTM, its ability to survive within macrophages, and its waxy outer membrane, effective antimicrobial treatment of NTM infections is difficult.

Due to the indolent nature of NTM infections, individuals may not seek care for months or years after infection.^8^ The official 2007 American Thoracic Society/Infectious Diseases Society of America (ATS/IDSA) guidelines for NTM treatment recommend initial treatment for MAC to be a cocktail therapy composed of at least three systemic antibiotics such as a macrolide (if susceptible), rifamycin, and ethambutol.^12^ On the other hand, the fast-growing *M. abscessus* are more likely to have antibiotic resistance and are initially treated with a combination of intravenous and oral antibiotics based on the strain’s susceptibilities.^13^

According to the 2007 ATS/IDSA guidelines, treatment for an NTM infection should be initiated if a patient presents with symptoms, relevant radiographic findings, and has confirmatory cultures.^13^ Treatment of a pulmonary MAC infection is recommended to be continued for at least 12 months after respiratory cultures convert to negative. Despite intensive treatment, culture conversion rates are often low, on average 55-65% of patients, with some studies reporting less than one third of patients clearing their infection.^12,14^ Meanwhile, *M. abscessus* infections lack the evidence to suggest a treatment length. The ATS/IDSA guidelines even suggests that pulmonary *M. abscessus* infections may be considered chronic infections.^13^ These chronic infections can hasten mortality, with one study of 167 patients with NTM pulmonary disease suggesting a survival estimate of 13.0 years for MAC infections compared to 4.6 years for other mycobaterial infections.^15^ The current dearth of established treatment options for NTM disease, and particularly for non-MAC and macrolide-resistant NTM infections, means that many patients would benefit from research into additional NTM antibiotics. Therefore, with advanced technology like high-throughput screening, it is crucial to discover new NTM antibiotics with better efficacy and tolerability.

Clofazimine (CFZ), which was first discovered in 1954 and marketed in 1969 by Novartis under the trade name Lamprene, is currently being considered for use in the treatment of NTM disease. CFZ is a bright red riminophenazine that is highly lipophilic and accumulates in the skin and macrophages. In addition to skin pigmentation, its main side effects include gastrointestinal upset and QT prolongation.^16^ It was first indicated for lepromatous leprosy but was quickly being replaced with more effective and less toxic drugs that came to market.^17^ Clinical interest in CFZ has revived in recent years in conjunction with its addition to the World Health Organization’s suggested treatment regimen for multidrug-resistant tuberculosis.^18^ CFZ’s exact mechanism of action is unknown, though the creation of reactive oxygen species and disruption of the cell membrane are proposed to be responsible for its antimicrobial activity.^17^ When taken up by an infected macrophage, CFZ seems to induce apoptosis of the macrophage thus inhibit mycobacterial growth.^19^ Due to its poor water solubility, CFZ has the tendency to crystallize within macrophages. These biocrystals have been suggested to play a role in CFZ’s anti-inflammatory effects.^20^ With a long half-life and potential post-antibiotic effect CFZ has broad bacteriostatic activity against gram positive bacteria. In addition, due to its high lipophilicity, CFZ penetrates human macrophages as well as the waxy mycobacterial cell wall.^17^ Furthermore, one study has suggested that clofazimine has shown some synergistic activity when combined with other antimicrobial agents such as aminoglycosides and macrolides in treating mycobacterial infections.^21^ Moreover, multiple studies have indicated *in vitro* susceptibility of several NTM species to CFZ, in addition to possible *in vitro* synergy with amikacin or clarithromycin against rapidly growing NTM.^22-24^ Taking all this into consideration, CFZ has become a promising candidate for treatment of chronic infections caused by certain NTM.

Given the lack of quality data from clinical trials, the 2007 IDSA guidelines have little to say on the appropriateness of CFZ use in treating an NTM infection.^13^ CFZ currently can only be obtained in the United States only through expanded access with an IRB-approved protocol.

In this retrospective review, we demonstrate how data from expanded access can be collected and analyzed from a single health system’s electronic medical record. The primary objective of this retrospective review was to determine the feasibility of collecting and analyzing data from expanded access drug utilization. The secondary objective was to generate hypotheses about the use of clofazimine in nontuberculous mycobacterial infections.

## Methods

### Patient identification

This was designed as a single-center, retrospective chart review, which was exempted from IRB oversight. Patients were identified through two methods: 1) using regulatory records of the expanded access cases and 2) conducting a data search in the health system’s electronic health record (EHR) for patients whose medication list contained clofazimine. The retrospective review included male and female patients of all races/ethnicities. Per inclusion criteria, Michigan Medicine patients had to have received at least one dose of CFZ before 30 June 2021 to be included in data analysis. Once patients were identified, direct identifiers were removed to retain patient confidentiality.

### Data Collection

Two of the authors independently collected data and resolved discrepancies through discussion. Data on all patients were collected through Michigan Medicine’s Electronic Medical Record Search Engine (EMERSE), which had access to notes, radiology, and pathology records since 1998.^25^ Data was collected until 30 June 2021 on each patient’s demographic (e.g., sex, DOB, weight, BMI (kg/m^2^), site of NTM infection, initial diagnosis date and NTM species as indicated in the notes, concomitant anti-mycobacterial medications, risk factors for NTM, initiation and end date (if applicable) of CFZ, CFZ dose, reasons for stopping CFZ, and any relevant symptoms, diagnostic scans (e.g., radiology, pathology), or cultures following initiation of CFZ. The patient’s medical records were then reviewed to determine pertinent clinical outcomes. The controllable search function of EMERSE enabled color-coded document highlighting and suggested synonyms to capture all incidences of a desired word. These synonyms included variations on a word’s spelling, alternative names, and abbreviations. As an example, synonyms of “clofazimine” included “clofazamine”, “Lamprene”, and “CFZ”, among others.

### Retrospective Review Population

A total of 55 patients were identified as having received clofazimine through expanded access records or medication lists. Of these patients, 11 were excluded as they had not initiated clofazimine (CFZ) treatment at the time of the retrospective review, and 5 additional patients were excluded due to ongoing treatment of fewer than 6 months, which was considered insufficient to show potential benefit. Two were excluded due to multiple concurrent NTM infections, and 1 was excluded due to discontinuation of treatment before 6 months that was not due to adverse events or negative culture. The remaining 36 patients were included in the analysis, with records analyzed between July 1, 2013, and June 30, 2021.

Patients were divided into subsets for analysis. The presence of one or more negative cultures was determined to be a sign of “effective” treatment, and patients who did or did not obtain this milestone were compared. Groups were also divided based on the causative agent of NTM infection – MAC or *M. abscessus*. The subset of patients infected with other mycobacterium species were not included in these comparisons except where noted.

### Statistical Analysis

Descriptive statistics were presented for categorical measures as frequencies (percent) and for continuous measures as medians (range). All univariable comparisons were performed using the Fisher’s exact test for categorical variables and the Wilcoxon rank sum test for continuous variables. A 2-tailed p-value of <0.05 was considered statistically significant. SAS 9.4 statistical software (Cary, North Carolina) was used for all analyses. Unless otherwise stated, all measures described have a nonsignificant association (p>0.05).

## Results

Analysis of the overall patient group showed a population in line with those previously reported in the literature.^8^ These reports are detailed in **Table 1**. The median age of patients at NTM diagnosis was 51.3 years, with a median age of 57.3 at the initiation of CFZ treatment. A median of 1.4 years elapsed between diagnosis and the decision to start CFZ. Most patients were female (64%), and the median BMI was 21.2 kg/m^2^. Seventy-two percent had a history of lung disease, with the most common diagnosis being asthma (28%), cystic fibrosis (22%), and chronic obstructive pulmonary disease (COPD) (17%). Fifty-two percent were former or current smokers. Thirty-nine percent had a history of either an immune-compromising condition or treatment.

**Table 1.** Demographic and Baseline Characteristics by Effectively Treated.

| Characteristic | Overall (n=36) | Yes (n= 19) | No (n=17) |
| --- | --- | --- | --- |
| Age (y) at NTM dx | 51.3 (12.1 to 91.3) | 52.9 (12.5 to 77.1) | 47.1 (12.1 to 91.3) |
| Age (y) at CFZ initiation | 57.3 (15.8 to 91.5) | 54.6 (16.3 to 80.9) | 57.9 (15.8 to 91.5) |
| Time (y) to CFZ initiation from NTM dx | 1.4 (0.0 to 21.6) | 0.5 (0.1 to 7.9) | 4.6 (0.0 to 21.6) |
| BMI (kg/m2)* (n=31) | 21.2 (13.5 to 28.5) | 21.8 (17.7 to 28.5) | 19.8 (13.5 to 28.5) |
| Female | 23 (64) | 11 (58) | 12 (71) |
| History of lung disease | 26 (72) | 11 (58) | 15 (88) |
| Cystic fibrosis | 8 (22) | 3 (16) | 5 (29) |
| Asthma | 10 (28) | 5 (26) | 5 (29) |
| COPD | 6 (17) | 1 (5) | 5 (29) |
| Other^ | 12 (33) | 5 (26) | 7 (41) |
| Former or current smoker* (n=31) | 16 (52) | 6 (40) | 10 (63) |
| Immunocompromised Status** | 14 (39) | 9 (47) | 5 (29) |
| NTM species |  |  |  |
| MAC | 17 (47) | 6 (32) | 11 (65) |
| M. abscessus | 13 (36) | 9 (47) | 4 (24) |
| Other | 6 (17) | 4 (21) | 2 (12) |
| Site of infection |  |  |  |
| Bone | 1 (3) | 1 (5) | 0 (0) |
| Disseminated | 4 (11) | 4 (21) | 0 (0) |
| Lung | 28 (78) | 12 (63) | 16 (94) |
| Skin | 3 (8) | 2 (11) | 1 (6) |
| Cough* (n=35) | 22 (63) | 10 (56) | 12 (71) |
| Fatigue* (n=35) | 5 (14) | 3 (17) | 2 (12) |
| Fever* (n=35) † | 1 (3) | 0 (0) | 1 (6) |
| Hemoptysis* (n=35) † | 2 (6) | 1 (6) | 1 (6) |
| Pain* (n=35) | 5 (14) | 5 (28) | 0 (0) |
| Phlegm* (n=35) | 18 (51) | 6 (33) | 12 (71) |
| Bronchiectasis* (n=33) | 15 (45) | 8 (44) | 7 (47) |
| Nodules* (n=33) | 20 (63) | 11 (65) | 9 (60) |
| Cavitary lesion* (n=33) | 9 (27) | 4 (22) | 5 (33) |

Values are median (range) or n (%).
\*Incomplete data resulted in a lower n
^Other lung diseases include pneumothorax, interstitial lung disease, ciliary dyskinesia, and alpha-1 antitrypsin deficiency
\*\*Immunocompromised status include active cancer/chemotherapy, multiple sclerosis, prednisone >20 mg/d, primary biliary cholangitis, CVID, IgG monoclonal gammopathy, and use of etanercept, methotrexate, and/or certolizumab

*M. avium* complex infections were the most common (47%), followed by *M. abscessus* (36%). The lung was the most common site of infection (78%), followed by disseminated disease (11%), and skin (8%). The most common symptoms reported at the initiation of treatment were cough (63%), excess sputum production (51%), fatigue (14%), and pain (14%). Nodules were found on radiographic imaging prior to CFZ treatment in 63%, bronchiectasis in 45%, and cavitary lesions in 27%. Overall, 53% of patients obtained a negative culture during treatment, while 47% did not.

Descriptive statistics showed small differences between the overall group, those who obtained at least one negative culture (“effectively treated,” n=19), and those who did not (“not effectively treated”, n=17). The median time from initial NTM diagnosis to treatment with CFZ was considerably longer (p=0.038) in patients who were not effectively treated (4.6 years) versus the effectively treated groups (0.5 years). The patients in the not effectively treated group were also more likely to have a history of lung disease (88%) compared to those who were in the effectively treated group (58%), which is exaggerated in the history of the individual conditions cystic fibrosis (29% versus 16%) and COPD (29% versus 5%). The most common species of infection was *M. abscessus* in the effectively treated group (47%) and MAC within the not effectively treated group (65%). The effectively treated group also contained 4 patients with other species that were not *M. avium* or *M. abscessus* (21%). Further results of these comparisons are available in **Table 1**.

To examine this difference, the overall patient population was divided into those with *M. abscessus* (n=13) and those with MAC infections (n=17). Full results are reported in **Table 2**. This revealed differences between the patients with each infection type. The median age in the *M. abscessus* group was younger at diagnosis (29.6 versus 47.1 years), younger at CFZ initiation (30.8 versus 56.8 years) and had less time between diagnosis and CFZ treatment (1.7 versus 3.7 years). While both groups had similar rates of previous lung disease (77% versus 76%), the *M. abscessus* group was marginally more likely to have had cystic fibrosis (38% vs. 18% for MAC) or asthma (31% vs. 18% for MAC), while a history of COPD was only found in the MAC group (35%, p=0.024). Similarly, there were former or current smokers in the MAC group (53%) versus the *M. abscessus* group (20%). Symptoms varied slightly, with somewhat more cough reported at the initiation of CFZ in the *M. abscessus* group (85% vs 59%), but somewhat less excess sputum production (46% vs 65%) and less fatigue (8% vs 24%). On radiographic imaging prior to starting treatment, the *M. abscessus* group was more likely to have bronchiectasis (75% vs 33%). The difference in the frequency of patients who were effectively treated with CFZ remained higher in the *M. abscessus* group (69%) over the MAC group (35%) but did not reach significance (p=0.139).

**Table 2.** Demographic and Baseline Characteristics by Species (*M. abscessus*/MAC only)

| Characteristic | Overall (n=30) | *M. abscessus* (n=13) | MAC (n=17) |
| --- | --- | --- | --- |
| Age (y) at NTM dx | 47.0 (12.1 to 91.3) | 29.6 (12.5 to 91.3) | 47.1 (12.1 to 77.1) |
| Age (y) at CFZ initiation | 54.1 (15.8 to 91.5) | 30.8 (16.3 to 91.5) | 56.8 (15.8 to 77.4) |
| Time (y) to CFZ initiation from NTM dx | 3.6 (0.2 to 21.6) | 1.7 (0.2 to 11.3) | 3.7 (0.3 to 21.6) |
| BMI (kg/m<sup>2</sup>)* (n=26) | 19.7 (13.5 to 28.5) | 22.0 (17.9 to 28.5) | 19.3 (13.5 to 26.8) |
| Female | 20 (67) | 8 (62) | 12 (71) |
| History of lung disease | 23 (77) | 10 (77) | 13 (76) |
| Cystic fibrosis | 8 (27) | 5 (38) | 3 (18) |
| Asthma | 7 (23) | 4 (31) | 3 (18) |
| COPD | 6 (20) | 0 (0) | 6 (35) |
| Other<sup>^</sup> | 12 (40) | 5 (38) | 7 (41) |
| Former or current smoker* (n=25) | 10 (40) | 2 (20) | 8 (53) |
| Immunocompromised Status** | 10 (33) | 4 (31) | 6 (35) |
| Effectively Treated | 15 (50) | 9 (69) | 6 (35) |
| Site of infection |  |  |  |
| Bone | 1 (3) | 1 (8) | 0 (0) |
| Disseminated | 3 (10) | 1 (8) | 2 (12) |
| Lung | 26 (87) | 11 (85) | 15 (88) |
| Cough | 21 (70) | 11 (85) | 10 (59) |
| Fatigue | 5 (17) | 1 (8) | 4 (24) |
| Fever | 1 (3) | 0 (0) | 1 (6) |
| Hemoptysis | 2 (7) | 1 (8) | 1 (6) |
| Pain | 2 (7) | 2 (15) | 0 (0) |
| Phlegm | 17 (57) | 6 (46) | 11 (65) |
| Bronchiectasis* (n=27) | 14 (52) | 9 (75) | 5 (33) |
| Cavitary lesion* (n=27) | 8 (30) | 4 (33) | 4 (27) |
| Nodules* (n=26) | 18 (69) | 8 (73) | 10 (67) |

Values are median (range) or n (%).
\*Incomplete data resulted in a lower n
<sup>^</sup>Other lung diseases include pneumothorax, interstitial lung disease, ciliary dyskinesia, and alpha-1 antitrypsin deficiency
\*\*Immunocompromised status include active cancer/chemotherapy, multiple sclerosis, prednisone >20 mg/d, primary biliary cholangitis, CVID, IgG monoclonal gammopathy, and use of etanercept, methotrexate, and/or certolizumab

The concomitant antibiotics prescribed in combination with CFZ are reported in **Table 3**. Note that more than one antibiotic could be used at the same time, though some antibiotics were more limited in their duration (e.g., cefoxitin was usually only administered for 8-9 weeks). In both the effectively treated and not effectively treated groups, the most commonly used antibiotics in combination with CFZ to target NTM were macrolide (74% and 65%) and IV or inhaled aminoglycosides (58% and 59%). The use of tetracycline was more common in the effectively treated group (37% versus 0%, p=0.008) while the use of rifamycin was less common (16% versus 53%, p=0.033).

**Table 3.** Concomitant Antibiotics by Effectively Treated.

| Characteristic | Overall (n=36) | Yes (n=19) | No (n=17) |
| --- | --- | --- | --- |
| Aminoglycoside | 21 (58) | 11 (58) | 10 (59) |
| Bedaquiline<sup>†</sup> | 2 (6) | 1 (5) | 1 (6) |
| Carbapenem<sup>†</sup> | 2 (6) | 1 (5) | 1 (6) |
| Cephalosporin<sup>†</sup> | 3 (8) | 3 (16) | 0 (0) |
| Ethambutol | 15 (42) | 6 (32) | 9 (53) |
| Fluoroquinolone<sup>†</sup> | 4 (11) | 2 (11) | 2 (12) |
| Macrolide | 25 (69) | 14 (74) | 11 (65) |
| Oxazolidinone | 5 (14) | 4 (21) | 1 (6) |
| Rifamycin | 12 (33) | 3 (16) | 9 (53) |
| Tetracycline | 7 (19) | 7 (37) | 0 (0) |
| Values are n (%). |  |  |  |
| <sup>†</sup> For antibiotics with <5 patients treated, Fisher's Exact tests were not conducted. |  |  |  |

In comparison, the concomitant antibiotics by common cause of NTM disease can be seen in **Table 4**. Aminoglycoside and macrolide drugs were the most commonly used among the group of patients with *M. abscessus* infections (69% and 62%). The most common antibiotics among the MAC group were ethambutol (82%), macrolide (76%), rifamycin (59%), and aminoglycoside (53%). Ethambutol and rifamycin were exclusively used in the MAC group (p<0.001 and p=0.001, respectively). Tetracycline was used by 38% of the *M. abscessus* group but none of the MAC group (p=0.009).These differences in antibiotic use are more likely a reflection of the NTM species spread among the groups than a reflection of possible synergism with CFZ.

**Table 4.** Concomitant Antibiotics by Common Cause of Species (*M. abscessus*/MAC only)

| Characteristic | Overall (n=30) | *M. abscessus* (n=13) | MAC (n=17) |
| --- | --- | --- | --- |
| Aminoglycoside | 18 (60) | 9 (69) | 9 (53) |
| Bedaquiline<sup>†</sup> | 2 (7) | 1 (8) | 1 (6) |
| Carbapenem<sup>†</sup> | 1 (3) | 1 (8) | 0 (0) |
| Cephalosporin<sup>†</sup> | 3 (10) | 3 (23) | 0 (0) |
| Ethambutol | 14 (47) | 0 (0) | 14 (82) |
| Fluoroquinolone<sup>†</sup> | 4 (13) | 0 (0) | 4 (24) |
| Macrolide | 21 (70) | 8 (62) | 13 (76) |
| Oxazolidinone<sup>†</sup> | 3 (10) | 2 (15) | 1 (6) |
| Rifamycin | 10 (33) | 0 (0) | 10 (59) |
| Tetracycline | 5 (17) | 5 (38) | 0 (0) |

Values are n (%).
<sup>†</sup> For antibiotics with <5 patients treated, Fisher's Exact tests were not conducted.

## Discussion

In the review of the data that were collected, the patient population that received CFZ at our health system has comparable characteristics to NTM patient populations described previously.^8^ Patients were majority female, had a history of lung disease including COPD, and had low-normal BMI. A large portion of patients had pulmonary NTM infections, with MAC representing the most common causative agent. Among the patients with lung infections, most presented with bronchiectasis and lung nodules at the time of initiation of CFZ, although it was unclear if this was a predisposing factor or a result of the infection.

In contrast to past epidemiological studies, this population had a higher-than-expected number of patients experiencing *M. abscessus* infections (36%), and the subset of patients with these infections differed in some important characteristics from the MAC patients. Although the differences did not reach statistical significance, the median age of *M. abscessus* patients was younger (29.6 versus 47.1 years), and the time between NTM diagnosis and initiation of CFZ treatment was less (1.7 versus 3.7 years). Although both sets had similar rates of lung infection and predisposing lung diseases, the *M. abscessus* patients were much more likely to have cystic fibrosis and much less likely to have COPD, which has been found in previous studies.^12-13^ They were also less likely to be former or current smokers, which similarly supports previous findings.^26^

More prominently, our data suggested that twice the rate of patients with *M. abscessus* infection experienced at least one negative culture than the patients with MAC (69% versus 35%), but this did not reach statistical significance. This result is biologically plausible, as antibiotics would be expected to be more effective on rapidly growing bacteria such as *M. abscessus* compared to slow-growing bacteria such as *M. avium*. However, the apparent difference may also be due to confounding factors such as the younger age of the *M. abscessus* patients, the shorter time to initiation of treatment, or the low number of patients with COPD. It is important to note that this finding is in conflict with an in-vitro study has specifically suggested that CFZ should be more effective in treating *M. avium* than in *M. abscessus*.^21^

Other studies have only been able to show the synergistic activities of CFZ when combined with other recommended antibiotics.^27^ Treatment with CFZ with at least two or more different antibiotics can possibly be beneficial in treating NTM infections without causing regrowth or bacterial resistance. This may also affect the outcomes of the patient population analyzed, as all patients had concomitant antibiotics.

### Retrospecitve Review Limitations

The primary objective of this retrospective review was to determine the feasibility of collecting and analyzing retrospective data from the use of drugs through expanded access. As the results show, retrospective data collection in these cases is possible but with some caveats.

Since data came from the electronic health record, the accuracy and completeness of the data were dependent on providers recording relevant information and on patient recall. The narrative format of progress notes exacerbated this ambiguity, as it required significant interpretation by the authors and often revealed inconsistent information (i.e., resolution of a symptom that had never been previously reported) that could not be converted into quantitative data. The attribution of symptoms was also often unclear, and with many patients experiencing NTM infection in the context of other pulmonary diseases or underlying conditions, it was very difficult even for a physician to distinguish the causality. The EMERSE search engine proved a powerful tool to resolve some of these concerns, as it was able to highlight symptoms across unstructured notes, but this was still prone to missing important information or requiring time-consuming interpretation and discussion.

Progress notes were not the only source of ambiguity. Active medication lists were often in conflict with the reported drug regimens and only rarely contained CFZ itself, as it is not an approved drug on the formulary. Medication start-dates appeared to be related more to prescription refills than the actual start of a treatment plan, wherein medications, which were used in the treatment extended over multiple years, would be repeated rather than updated. For patients who received some of their care outside the institution, this information was not populated in a consistent manner. Hence, all of these factors limited the reliability of data and increased the complexity of data collection.

This was a small, single-site feasibility retrospective data review, which necessarily limits the generalizability of any conclusions. The structure of electronic health record systems, as well as the institutional culture around what progress notes contain, may affect how feasible this type of data collection is at any given site. Within the studied institution, the structure of these records limited the ability to draw conclusions about symptomology or specific regimens. Data review was also retrospective, which further limits the ability to collect specific endpoints.

Further, the small sample size of 36 patients meant that few of the results reached statistical significance. Patients at this site may be fundamentally different from NTM patients nationwide, both because the institution reviewed is a large academic medical center and because there is known variability in epidemiological characteristics by location. The findings are intended to be hypothesis-generating but warrant future study of a sufficient size and diversity.

## Conclusion

Collecting real-world data for drugs obtained through expanded access pathways is feasible but may be complicated by the structure of electronic health systems, the content of progress notes, and the need for interpretation of narrative data. This data can provide observations, such as the higher rate of successful treatment among *M. abscessus* patients as compared to MAC patients receiving clofazimine, that may impact the clinical use of this drug. However, given the retrospective nature of the collection and the inconsistencies noted in the data, future studies are required to validate these findings.

## Data Availability

All data produced in the present study are available upon reasonable request to the authors.

